# Forecasting Influenza-Like Illness (ILI) during the COVID-19 Pandemic

**DOI:** 10.1101/2022.10.27.22281617

**Authors:** Stephen D. Turner, Chris Hulme-Lowe, VP Nagraj

## Abstract

Near-term probabilistic forecasts for infectious diseases such as COVID-19 and influenza play an important role in public health communication and policymaking. From 2013-2019, the FluSight challenge run by the Centers for Disease Control and Prevention invited researchers to develop and submit forecasts using influenza-like illness (ILI) as a measure of influenza burden. Here we examine how several statistical models and an autoregressive neural network model perform for forecasting ILI during the COVID-19 pandemic, where historical patterns of ILI were highly disrupted. We find that the autoregressive neural network model which forecasted ILI well pre-COVID still performs well for some locations and forecast horizons, but its performance is highly variable, and performs poorly in many cases. We found that a simple exponential smoothing statistical model is in the top half of ranked models we evaluated nearly 75% of the time. Our results suggest that even simple statistical models may perform as well as or better than more complex machine learning models for forecasting ILI during the COVID-19 pandemic. We also created an ensemble model from the limited set of time series forecast models we created here. The limited ensemble model was rarely the best or the worst performing model compared to the rest of the models assessed, confirming previous observations from other infectious disease forecasting efforts on the less variable and generally favorable performance of ensemble forecasts. Our results support previous findings that no single modeling approach outperforms all other models across all locations, time points, and forecast horizons, and that ensemble forecasting consortia such as the COVID-19 Forecast Hub and FluSight continue to serve valuable roles in collecting, aggregating, and ensembling forecasts using fundamentally disparate modeling strategies.

## 1 Introduction

Collaborative infectious disease forecasting efforts are critical to an informed public health response. Over the past decade, several consortia have emerged to provide high-quality, timely predictions of disease spread to forecast consumers in the United States (Reich, McGowan, et al. 2019; Cramer et al. 2022). In addition to delivering operational epidemiological forecasts, these consortia have contributed to a growing literature about the effectiveness of various infectious disease modeling approaches. One of the most well established epidemic forecasting consortia is the community of forecasters participating in the Center for Disease Control and Prevention (CDC) FluSight initiative. FluSight was initially conceived in 2013, with the goal of encouraging academic and industry researchers to develop methods to forecast the timing, peak, and intensity of the flu season in the United States (CDC 2019). Participants in the FluSight challenge have implemented and assessed a wide variety of methods for forecasting influenza-like illness (ILI), including compartmental models, statistical (e.g., time series) models, and machine learning approaches. Infectious disease modelers have demonstrated the utility of time series methods for forecasting ILI, and the performance of specific approaches such as autoregressive integrated moving average (ARIMA) and autoregressive neural network (AR-NN) has been studied previously (Kandula and Shaman 2019a; Meng, Huang, and Kong 2022; Tsan et al. 2022).

The onset of the COVID-19 pandemic disrupted many aspects of daily life globally and shifted infectious disease modeling focus away from influenza. Around the world, reported levels of flu activity dropped precipitously beginning in 2020 (Zipfel, Colizza, and Bansal 2021). Some researchers have postulated that non-pharmaceutical interventions implemented for COVID-19 have helped curb the spread of other respiratory pathogens, including flu (Chan et al. 2020; Feng et al. 2021; Sun et al. 2022; Youssef et al. 2022). At least one study further acknowledges that the decline in ILI activity may be related to changes in healthcare-seeking or the excess burden on health systems from COVID-19 (Jayaram et al. 2022). Whatever the exact mechanism, the ILI signal has been dramatically interrupted by COVID-19.

Although ILI has undoubtedly been impacted by the spread and response to COVID-19, the signal continues to be reported as a surveillance endpoint in the United States. Methods for forecasting ILI continue to be studied (McAndrew and Reich 2021). We suggest that, given the disruption to ILI by COVID-19, the performance of methodology developed pre-pandemic is worth revisiting. In particular, time series approaches for ILI forecasting may exhibit different performance when trained with data collected since 2020. Here we present the results of an analysis of forecasts of near-term ILI targets in the United States, using a selection of time series models trained on data collected during the COVID-19 pandemic. Our aim is to evaluate these methods side-by-side in order to inform future ILI forecasting efforts that may encounter continued interference to the ILI indicator from COVID-19.

## 2 Methods

### 2.1 Data sources

The Outpatient Influenza-like Illness Surveillance Network (ILINet) aggregates and distributes summaries of ILI activity (i.e., percentage of outpatient visits with fever plus cough or sore throat) nationally, regionally, by state. CDC FluView provides an interface to retrieve current and historical ILINet data down to the state level. We retrieved ILI signal from CDC FluView from October 2010 (the beginning of the 2010/2011 season) through June 2022 (the end of the 2021/2022 season). The flu season is generally considered to run from fall through late spring, however ILI data is reported during the summer months as well. We collected ILI data for all weeks available (including weeks in the summer outside the flu season) for the U.S. as a whole and for each individual state. Florida was the only location excluded due to inconsistent data availability across the window of time we studied. Note that we used “weighted” value of ILI, which takes into account state population. For any given state, the weighted and unweighted ILI will be equal. However, when aggregated (e.g., nationally) the weighted ILI balances the activity indicator by population.

We created one to four week ahead forecasts from all weekly forecast dates ranging between February 7, 2021 and May 22, 2022. We established two different time windows for ILI training data: (1) a “long” data set encompassing all weeks from October 2010 through the retrospective date when the forecast was made (the “forecast date”) and (2) a “short” data set consisting of all data from January 2020 through the forecast date. The “short” training series was used to assess how models performed when excluding data from previous seasons when COVID-19 was not circulating in the U.S.

### 2.2 Forecasting methods

We used four different time series models to generate ILI forecasts: autoregressive integrated moving average (ARIMA), seasonal autoregressive integrated moving average (SARIMA), exponential smoothing (ETS), and autoregressive neural networks (AR-NN). All models were fit using the fable (O’Hara-Wild, Hyndman, and Wang 2021a) and fabletools (O’Hara-Wild, Hyndman, and Wang 2021b) packages in R (R Core Team 2022), in part using infrastructure described in (Nagraj et al. 2022). Models were trained using the ILI data described above, with long and short training sets used for all models. In some cases, the parameterization of models yielded alternative approaches from the same model family (for example, see “unrestricted” and “restricted” ARIMA below). In total, we fit 12 different combinations of models and training data. To generate forecasts, we iteratively masked data in the testing window and stepped through each week to generate one to four week ahead forecasts. As each week in the testing window was eclipsed, the models were retrained to include the additional week of training data. For example, we forecasted horizons beginning at the week of February 7, 2021 using models trained on short and long ILI data, then we iterated to the next forecast week (February 14, 2021) using the short and long training data augmented with observations from the week of February 7, 2021. This procedure was repeated to advance through all weekly forecast dates ending in May 22, 2022.

#### 2.2.1 Autoregressive integrated moving average (ARIMA)

One of the approaches we evaluated was an auto ARIMA modeling technique. ARIMA models are one of the most widely used approaches for forecasting time-dependent data, and have been used to model ILI in previous studies (R. Hyndman and Athanasopoulos 2021) (Kandula and Shaman 2019b). Briefly, an ARIMA model is specified by three parameters: the number of lagged time points to include (*p*), the number of differences to take of the observed data (*d*), and the order of the moving average (*q*). Two different approaches were taken to fitting the ARIMA model. The first closely mimics the approach used by Kandula and Shaman (2019b) in which the *p, d*, and *q* parameters were selected from an “unrestricted” space to minimize the corrected Akaike information criterion (AICc) (R. J. Hyndman and Khandakar 2008). The other approach selects the parameters and hyperparameters to minimize the AICc, but uses a “restricted” space such that *p* ∈ (0, 1, 2), *d* ∈ (0, 1), and *q* ∈ (0, 1, 2, 3, 4). We previously found that the restricted ARIMA approach worked well for estimating weekly COVID-19 incidence (Nagraj et al. 2021).

The ARIMA model described above is predicated on the idea that future observations of a variable are predicted by past observations of the same variable (R. Hyndman and Athanasopoulos 2021). It does not, however, explicitly account for seasonal variation in the observations, and seasonal fluctuation of ILI is well established (Kandula and Shaman 2019b). A generalized seasonal form of the ARIMA model, known as the SARIMA model, is also commonly used in predicting time dependent variables in situations where there are regular, predictable fluctuations in the observations. As with the non-seasonal ARIMA models, the SARIMA models parameters and hyperparameters were selected to minimize the AICc with no restrictions on the parameter spaces.

### 2.2.2 Exponential smoothing (ETS)

Like the ARIMA model described above, the exponential smoothing method for modeling a time series with error, trend, and seasonality (ETS) is frequently used to predict future observations from past data. Predictions are made by taking a weighted average of past observations of the same variable, where the weights decay exponentially as the observations become further removed from the present. As with the ARIMA and SARIMA models described above, ETS models can be modified to explicitly account for global trends and/or seasonal variation in the variable being predicted, making them a very flexible tool for forecasters. The rate at which the weights applied to the previous observations decrease can be controlled by a smoothing parameter during model fitting. Larger smoothing parameter values result in more weight being given to the most recent observations and the weights assigned to other values rapidly decreasing. Small values of the smoothing parameter, conversely, give relatively less weight to the most recent observations (although, they still receive greater weight than older observations) and a slower decay of the weights assigned to more temporally distant observations. In addition to the smoothing parameter, ETS models require that the initial state of the model, usually denoted ℓ_*o*_, be specified. Both the model’s initial state and its smoothing parameter are estimated from the observed data by maximizing log-likelihood.

### 2.2.3 Autoregressive neural networks

Recent research has demonstrated the utility of autoregressive neural network models (AR-NN) for forecasting ILI (Kandula and Shaman 2019a). The intuition behind this model is that by using lagged values of a variable as inputs mapped to a single hidden layer in a neural network, it is possible to model a non-linear autoregressive process, such as ILI intensity. The neural network framework allows for additional hidden layers to be added as needed. Following the methods described in Kandula and Shaman (2019a), we set the number of previous seasons to consider *P* to 1, the number of hidden layers to use *k* to 1, and then automatically selected the number of lagged input (*p*) to minimize the AICc. As with the other models, this optimization was performed using the fable R package.

### 2.2.4 Baseline and ensemble models

For comparison, we also implemented a “baseline” model, which is simply a naive forecast setting the value of all forecasts to be the value of the last observation. We also created an ensemble of models (excluding the baseline) by taking an equally weighted average of forecasts from all models at each quantile level.

### 2.3 Model evaluation

Assessing the performance of epidemiological models is a topic that has received considerable attention in the literature around the FluSight competition (Bracher 2019; Reich, Osthus, et al. 2019). At present, many infectious disease forecasters are using weighted interval scores (WIS; Bracher et al. (2021)) to evaluate consortia forecasts. The WIS approximates the continuous ranked probability score (CRPS) while penalizing for over- and under-prediction. WIS was proposed as an alternative to metrics such as the logarithmic score and CRPS, which are limited when the forecasts are prepared in interval format. Both the logarithmic score and the CRPS assume full knowledge of the distribution of the predictive distributions. While these can be approximated with an interval format prediction by computing many intervals, these approximations break down when the predictions fall into the tails of the predictive distributions. We calculated the WIS using the `evalcast R` package (McDonald et al. 2021).

We also calculated the relative weighted interval score (rWIS) for any given location, forecast date, and horizon, relative to the WIS of the naive baseline model described above. Following (Cramer et al. 2022) we calculated the standardized rank score for each model for each location - forecast date - horizon observation, which falls between 0 and 1 for every forecasted observation. For *i* models, the model with the best (lowest) rWIS received a rank of 1, and the worst received a rank of *n*_*i*_, and the standardized rank was rescaled between 0 and 1 where 0 corresponded to the worst rank and 1 to the best.

## 3 Results

For each forecast week, ILI forecasts for 50 locations using all of the models trained on both data sets and four different prediction horizons, resulted in 27,200 retrospective forecasts. Model performance was evaluated using the the rWIS (relative to the baseline model). We assessed model performance by examining rWIS medians, distributions, and standardized ranks across forecast dates and horizons.

### 3.1 rWIS median and distribution

Table 1 shows the median rWIS for national and state targets across forecast dates and horizons. Figure 1 shows the distribution of rWIS across forecast dates and horizons. In both Figure 1 and Table 1 all included states are lumped together in the “States” column or panel, with national-level data shown separately.

**Table 1:**
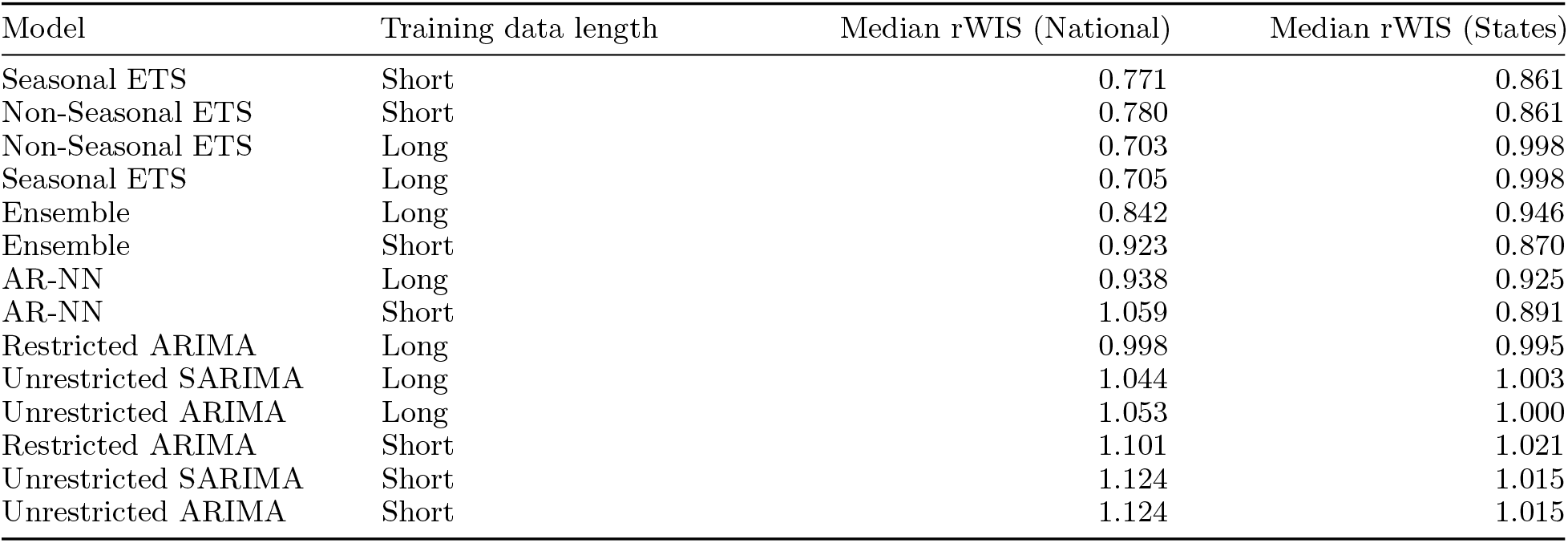
Median rWIS for national and state targets for each model and training data length combination.

| Model | Training data length | Median rWIS (National) | Median rWIS (States) |
| --- | --- | --- | --- |
| Seasonal ETS | Short | 0.771 | 0.861 |
| Non-Seasonal ETS | Short | 0.780 | 0.861 |
| Non-Seasonal ETS | Long | 0.703 | 0.998 |
| Seasonal ETS | Long | 0.705 | 0.998 |
| Ensemble | Long | 0.842 | 0.946 |
| Ensemble | Short | 0.923 | 0.870 |
| AR-NN | Long | 0.938 | 0.925 |
| AR-NN | Short | 1.059 | 0.891 |
| Restricted ARIMA | Long | 0.998 | 0.995 |
| Unrestricted SARIMA | Long | 1.044 | 1.003 |
| Unrestricted ARIMA | Long | 1.053 | 1.000 |
| Restricted ARIMA | Short | 1.101 | 1.021 |
| Unrestricted SARIMA | Short | 1.124 | 1.015 |
| Unrestricted ARIMA | Short | 1.124 | 1.015 |

**Figure 1:**
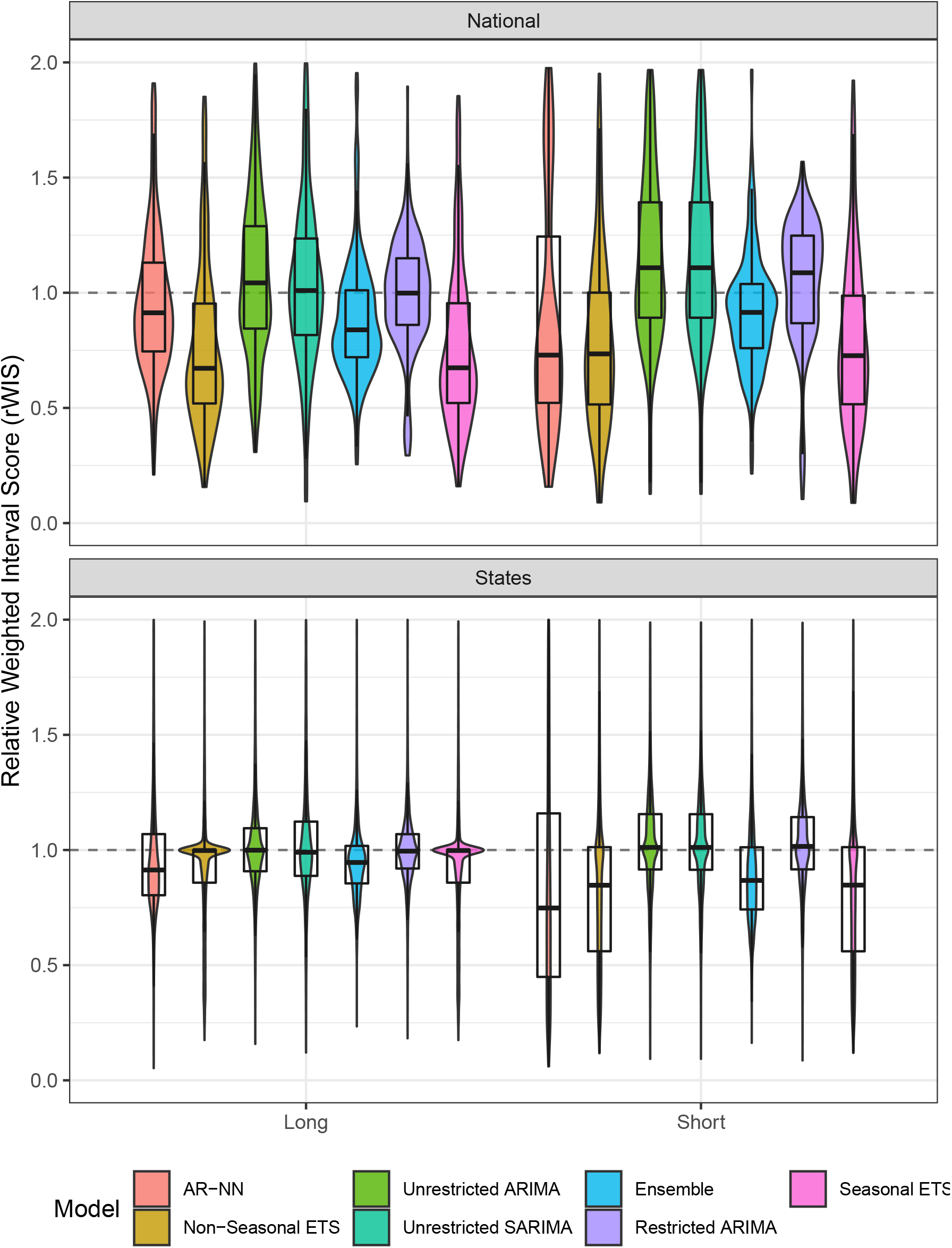
rWIS distributions for each model and training data length for National and State targets across all forecast dates and week-ahead horizons. Distribution is illustrated by a density polygon with a boxplot overlay. rWIS was truncated at 2.0 to aid in visualization, even though some models had extremely high rWIS values for some observations.

For national-level data, these results show that for both the long and short training data series, the ETS models had the lowest rWIS, the ARIMA formulations had the highest rWIS, with the ensemble and AR-NN models between (Figure 1, top panel). Of note, the AR-NN model had extremely variable performance when using the short training data. For states, the distributions of rWIS across forecast dates and horizons was highly variable. In general, the ETS models performed better on the short and long time series training data, and there was little difference in rWIS across model types using the long training data for states.

### 3.2 rWIS rank distribution

As was previously described in Cramer et al. (2022) for COVID-19 forecast models, we calculated the standardized rank for each model + training data length across locations, forecast dates, and horizons. Figure 2 shows these results, separately for national and state targets. These results show that model accuracy rankings are highly variable. For instance, while AR-NN (Short) model had a standardized rank of >0.9 for nearly 25% the targets forecasted, its performance was highly variable, as can be seen with the strongly bimodal distribution of standardized ranks for the AR-NN (Short) model and the long tail of poor performance with the AR-NN (Long) model in Figure 2. The simple exponential smoothing (seasonal or nonseasonal ETS with the short training series) was the only model that had a standardized rank of >0.5 for over 75% of the state target observations forecasted. It is also noteworthy here that the ensemble models (ensembled from individual models using either the short or the long training series) has less variable performance (the shape of the standardized rank distribution is roughly normal-shaped). The ensemble model seldom had the highest standardized rWIS rank, but was likewise rarely in the lowest standardized rWIS ranks.

**Figure 2:**
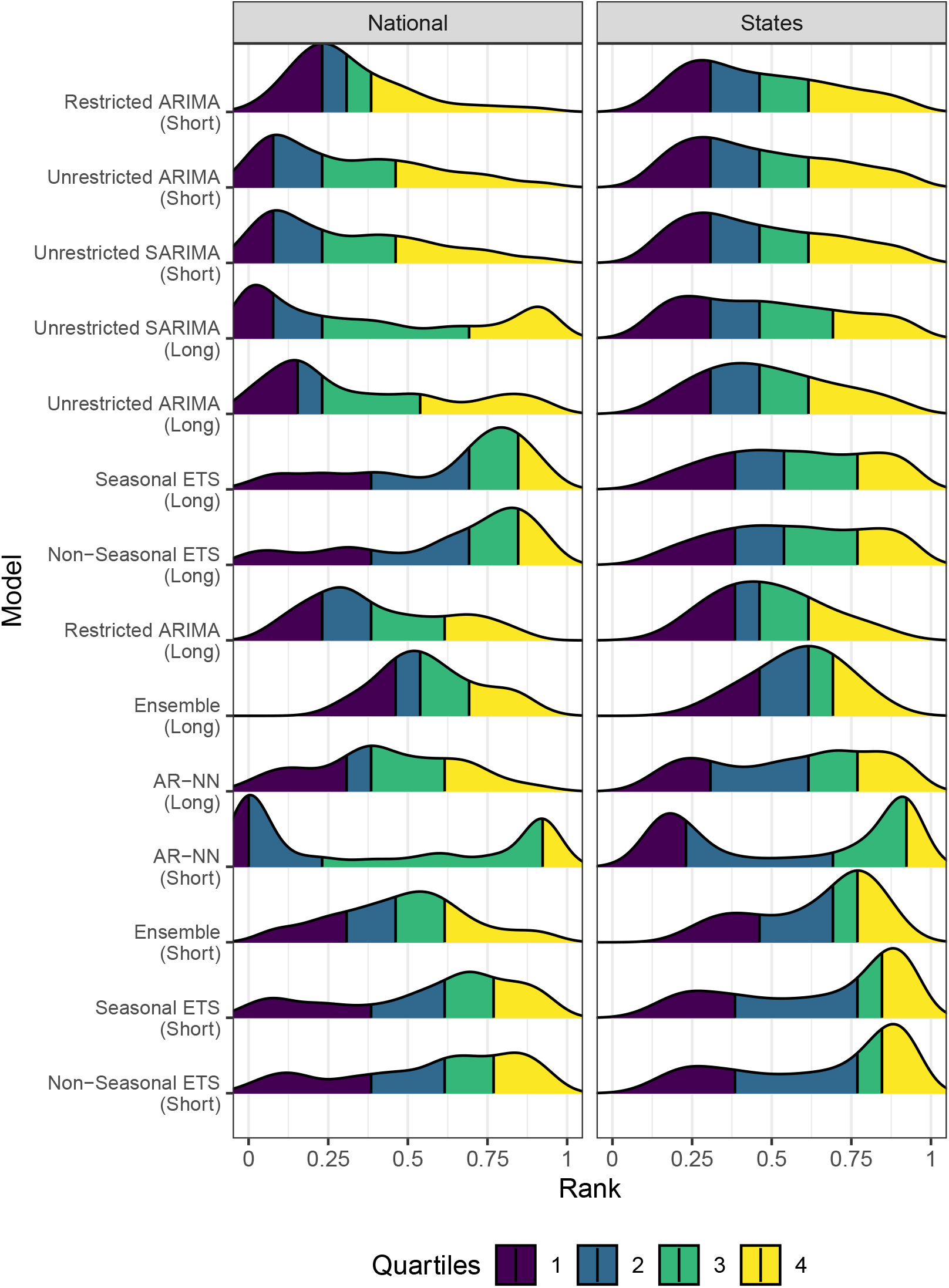
Standardized rWIS rank distribution for each model (and training data length) across locations, forecast dates, and horizons. A standardized rank of 1 indicates that the model had the lowest (best) rWIS across locations, targets, and forecast horizons, and a standardized rank of 0 indicates that the model had the worst (highest) rWIS. The quartiles of each model’s distribution of standardized ranks are shown by different colors with yellow indicating the top quartile and indigo representing the bottom quartile of the distribution. Models are ordered by model with the lowest median rWIS on the bottom to the highest median rWIS on the top, averaged across states and national targets.

## 4 Discussion

Previous research has shown that time series models can effectively be implemented to forecast ILI. In particular, autoregressive neural network (AR-NN) models when appropriately tuned can outperform statistical time series models such as ARIMA (Kandula and Shaman 2019a). When examining model performance for forecasting ILI during the COVID-19 pandemic using the lowest median rWIS as a heuristic for selecting the best-performing model, the results we present here support previous findings that neural network models are still a valuable tool for forecasting ILI. However, while the AR-NN model was a top performing model (very high standardized rWIS rank) for a portion of forecasts created here, its performance was highly variable, with extremely poor performance in some scenarios regardless of training data length. The results presented here suggest that when considering distributions of rWIS aggregated across locations, dates, and forecast horizons, traditional statistical approaches, even a simple exponential smoothing model, may have consistently better performance for forecasting the ILI signal observed with COVID-19 circulating.

This study has notable strengths and limitations. The analysis presented here includes ILI forecasts down to the state level. Much of the existing literature around ILI forecasting has focused on lower geographic resolution (i.e., HHS regions). Our analysis provides an informative view of the variability in forecasters across state-level targets. However, we acknowledge that the study design may limit generalizability of these results somewhat. At the time of publication of this manuscript there have only been two influenza seasons (2020-2021 and 2021-2022) that have occurred during the COVID-19 pandemic. As the pandemic has subsided in 2022 after successive waves of SARS-CoV-2 Omicron subvariants, pandemic restrictions have largely been lifted worldwide, and pre-pandemic behaviors are returning. It remains unclear whether the generally high performance of simple statistical models for forecasting ILI we observe here will remain durable. However, future SARS-CoV-2 variants may result in disruption to ILI measures in future flu seasons, and statistical time series approaches that do not rely on pre-pandemic training data may be useful for years to come.

Our results are consistent with high-level “no free lunch” observations of consortium forecasting efforts like FluSight and the COVID-19 Forecast Hub, in that no single model outperforms all other models for all locations, time points, and forecast horizons. Figure 2 shows this most clearly. Cramer et al. (2022) showed very similar results with COVID-19 in which all models showed large variability in relative skill, with each model having observations for which it had the lowest (best) rWIS, and with the ensemble model having less variable and favorable performance globally. Our ensemble model differs from that presented in Cramer et al. (2022) for COVID-19 in that our model *only* consists of time series models, and lacks the model diversity of the COVIDhub and FluSight ensembles, which include compartmental models, agent based models, human judgment forecasts, and time series approaches.

## Data Availability

Data used in this study are available via CDC FluView, and can be accessed programmatically and analyzed with code available at https://github.com/signaturescience/fiphde

## 5 Acknowledgements

This work was supported in part by a subaward to Signature Science, LLC from the Council of State and Territorial Epidemiologists (CSTE) via the Centers for Disease Control (CDC) Cooperative Agreement No. NU38OT000297.

